# Identifying Hepatocellular Carcinoma from imaging reports using natural language processing to facilitate data extraction from electronic patient records

**DOI:** 10.1101/2022.08.23.22279119

**Authors:** Tingyan Wang, Ben Glampson, Luca Mercuri, Dimitri Papadimitriou, Christopher R Jones, David A Smith, Hizni Salih, Cori Campbell, Oliver Freeman, Steve Harris, Kinga A Várnai, Gail Roadknight, Stephanie Little, Theresa Noble, Kerrie Woods, Philippa C Matthews, NIHR Health Informatics Collaborative Viral Hepatitis Theme Consortium, Jim Davies, Graham S Cooke, Eleanor Barnes

**Author notes:** Corresponding authors. The Peter Medawar Building for Pathogen Research, South Parks Road, Oxford, OX1 3SY, UK. Joint last authors, represents equal contribution (Jim Davies, Graham S Cooke, Eleanor Barnes). NIHR Health Informatics Collaborative (HIC) Viral Hepatitis Theme Consortium: Eleanor Barnes, Jim Davies, Kerrie Woods, Philippa C Matthews, William Gelson, Graham S Cooke, Salim I Khakoo, Eleni Nastouli, Alexander J. Stockdale, Stephen Ryder, Ahmed Elsharkawy, Douglas Macdonald, Kinga A Várnai, Tingyan Wang, Cori Campbell, Gail Roadknight, Stephanie Little, Ben Glampson, Dimitri Papadimitriou, Luca Mercuri, Christopher R Jones, Jakub Jaworski, Afzal Chaudhry, Vince Taylor, Cai Davis, Josune Olza, Florina Borca, Hang Phan, Frazer Warricker, Louise English, Luis Romão, David Ramlakhan, Stacy Todd, Heather Rogers, Karl McIntyre, Andrew Frankland, Steve Harris, Theresa Noble, David A Smith, Oliver Freeman, Sarah Montague, Irene Juurlink, Ruth Norris, George Tilston.

## Abstract

**Background:** The National Institute for Health Research Health Informatics Collaborative (NIHR HIC) viral hepatitis theme is working to overcome governance and data challenges to collate routine clinical data from electronic patients records from multiple UK hospital sites for translational research. The development of hepatocellular carcinoma (HCC) is a critical outcome for patients with viral hepatitis with the drivers of cancer transformation poorly understood.

**Objective:** This study aims to develop a natural language processing (NLP) algorithm for automatic HCC identification from imaging reports to facilitate studies into HCC.

**Methods:** 1140 imaging reports were retrieved from the NIHR HIC viral hepatitis research database v1.0. These reports were from two sites, one used for method development (site 1) and the other for validation (site 2). Reports were initially manually annotated as binary classes (HCC vs. non-HCC). We designed inference rules for recognising HCC presence, wherein medical terms for eligibility criteria of HCC were determined by domain experts. A rule-based NLP algorithm with five submodules (regular expressions of medical terms, terms recognition, negation detection, sentence tagging, and report label generation) was developed and iteratively tuned.

**Results:** Our rule-based algorithm achieves an accuracy of 99.85% (sensitivity: 90%, specificity: 100%) for identifying HCC on the development set and 99.59% (sensitivity: 100%, specificity: 99.58%) on the validation set. This method outperforms several off-the-shelf models on HCC identification including “machine learning based” and “deep learning based” text classifiers in achieving significantly higher sensitivity.

**Conclusion:** Our rule-based NLP method gives high sensitivity and high specificity for HCC identification, even from imbalanced datasets with a small number positive cases, and can be used to rapidly screen imaging reports, at large-scale to facilitate epidemiological and clinical studies into HCC.

**Statement of Significance:** 

**Problem:** Establishing a cohort of hepatocellular carcinoma (HCC) from imaging reports via manual review requires advanced clinical knowledge and is costly, time consuming, impractical when performed on a large scale.

**What is Already Known:** Although some studies have applied natural language processing (NLP) techniques to facilitate identifying HCC information from narrative medical data, the proposed methods based on a pre-selection by diagnosis codes, or subject to certain standard templates, have limitations in application.

**What This Paper Adds:** We have developed a hierarchical rule-based NLP method for automatic identification of HCC that uses diagnostic concepts and tumour feature representations that suggest an HCC diagnosis to form reference rules, accounts for differing linguistic styles within reports, and embeds a data pre-processing module that can be configured and customised for different reporting formats. In doing so we have overcome major challenges including the analysis of imbalanced data (inherent in clinical records) and lack of existing unified reporting standards.

## Background

Primary liver cancer (of which the vast majority are hepatocellular carcinoma; HCC) is the sixth most common cancer and the fourth leading cause of cancer-related mortality globally (1). There were approximately 0.9 million incident cases of HCC in 2020 and this will rise to an estimated ∼1.4 million in 2040 (2). HCC typically develops in the setting of chronic liver disease; chronic infection with hepatitis B or C virus (HBV or HCV), alcohol and metabolic syndromes are the most frequent HCC risk factors (3). However, most patients with HCC are diagnosed in the late stages of the disease when treatment options are limited (4), leading to poor outcomes (∼12% 5-year survival rate for these late-stage HCC patients (5)).

Imaging investigations are currently crucial in HCC surveillance and diagnosis as there are no sensitive tumour biomarkers for accurate early HCC detection (6). The European Association for the Study of the Liver (EASL) clinical guidelines (7) strongly recommend that HCC surveillance using abdominal ultrasound is performed every 6 months in populations at high-risk of HCC. Imaging modalities including multiphasic computed tomography (CT) or magnetic resonance imaging (MRI), usually with intravenous contrast injection, are non-invasive methods currently recommended in clinical guidelines for HCC diagnosis in patients with liver cirrhosis —usually where liver nodules have first been detected by ultrasound (7-10). Imaging reports are typically generated, in non-standardised narrative format by radiologist, after reviewing the scans (11). These reports typically contain vital information about the presence or absence, and stage of HCC.

Despite their potential utility, imaging reports exist mainly as free-text clinical narrative, which are difficult to query for secondary use. The potential utility of accurate automatic interpretation of these reports is enormous and includes clinical use in audit, diagnostic reporting and dissemination, financial management and planning of health service provision. Additionally these reports could be used for large national research programs, such as NIHR HIC, that seek to address important research questions for clinical benefit including HCC epidemiology, risk factors, biomarkers for early detection and clinical outcomes (12). Establishing an HCC cohort from unstructured imaging report data via manual review is costly, time consuming, impractical and requires advanced clinical knowledge when performed on a large scale (13). In the United States, the American College of Radiology (ACR) published the Liver Imaging Reporting and Data System (LI-RADS) to standardise the reporting of various imaging modalities for HCC (8), with each report assigned to a category. This means that these reports are relatively structured for easy for data query (14). However, this system has not yet been widely adopted in the UK, and it is currently impossible to gather even simple information, like HCC diagnosis from imaging reports in an automated fashion. There is a clear need to leverage computational approaches for extracting information such as HCC diagnosis from unstructured imaging reports.

The classification of HCC using imaging reports requires extracting relevant words and phrases that define an HCC diagnosis from these free-text reports, for which natural language processing (NLP) techniques provide promising automatic solutions. NLP is a range of computational techniques for analysing and representing naturally occurring texts at different levels of linguistic analysis for the purpose of accomplish human-like language processing for a range of tasks or applications. Text classification leveraging NLP is an attractive method that may be used to facilitate the development of precisely phenotyped clinical cohorts using free-text medical records (13, 19, 20). Over recent years, machine learning (ML) or deep learning (DL) based NLP models have been shown to be effective in text classification tasks (15, 16); however, in the medical domain there is often a scarcity of large amounts of manually labelled data that is required for the robust training of such models. More recently, advanced NLP techniques, one of which uses pre-trained language models based on transformers have been developed (17), for which minimal annotated data is required to fine-tune the model for specific downstream NLP tasks. However, it is still difficult to leverage the existing pre-trained models for text classification where the data is highly imbalanced such that there is a significantly lower proportion of imaging reports with HCC cases compared to those that do not suggest a HCC diagnosis. To overcome the challenges of small sample size of labelled data and high-degree imbalance of data, in this study we propose to develop a rule-based NLP method to classify imaging reports for HCC identification, as diagnosis information extraction based on HCC inference rules does not require large amounts of manually-annotated data as well as can handle the imbalance issue by direct keywords recognition and matching.

Previous studies have applied NLP techniques including ML or rule-based methods or hybrid approaches into HCC-related medical text to extract standardised descriptions of anatomical findings, annotate tumour characteristics, or track the tumour statuses based on reports from patients with HCC (12, 22-24). Other studies have focused on classifying imaging reports to predict downstream radiology resource utilisation in patients undergoing HCC surveillance (25). However, there are few studies (13,14) addressing the same task as this study that focused on how to identify information that defines an HCC diagnosis from imaging reports. One study conducted text classification for HCC identification using imaging reports that were pre-selected by international classification of disease (ICD) codes (13), which inherently does not address the high imbalance issue of imaging reports, nor consider the misclassification issue by diagnosis codes. Another study used a large bank of standard LI-RADS formatted imaging reports as annotated data to train models (14), which is not applicable in the region/ countries that do not widely use such template system for reporting imaging examination findings. Therefore, we set out to develop a rule-based NLP method which (a) utilises both diagnostic concepts and tumour feature representations that suggest an HCC diagnosis to form reference rules, (b) takes into account linguistic styles in different sections of imaging reports and the matching priority of medical terms, and (c) embeds data preprocessing steps that can be configured and customised for different reporting formats. Given these, our method will be capable of classifying HCC using imaging reports in real-world datasets that are being drawn automatically from operational systems (i.e., without pre-selection by diagnosis codes), as well as can be applied in imaging reports with different reporting styles, rather than being subject to certain templates e.g., LI-RADS reporting standard.

## Methods and Materials

### Data sources

Imaging reports used in this study were collected from patients with HBV, HCV, or HEV and stored in the central data repository of the NIHR HIC viral hepatitis theme, for which the data collection process has been detailed previously (15, 16). The imaging reports for this study cover various imaging modalities, including CT, MRI, and ultrasound, and were originally from two different sites: Oxford University Hospitals NHS Foundation Trust (site 1) and Imperial College Healthcare NHS Trust (site 2). Imaging reports were manually labelled (i.e., assigning a binary label (‘yes’ or ‘no’) to an imaging report to indicate whether there is HCC presence in that imaging examination). We then selected imaging reports that have been manually labelled to develop and validate a rule-based method.

### Rule-based NLP model

The framework of the proposed rule-based NLP method for HCC identification from imaging reports is shown in Figure 1, including text preprocessing, HCC inference rules, rule-based algorithm, and iterative development.

**Figure 1.**
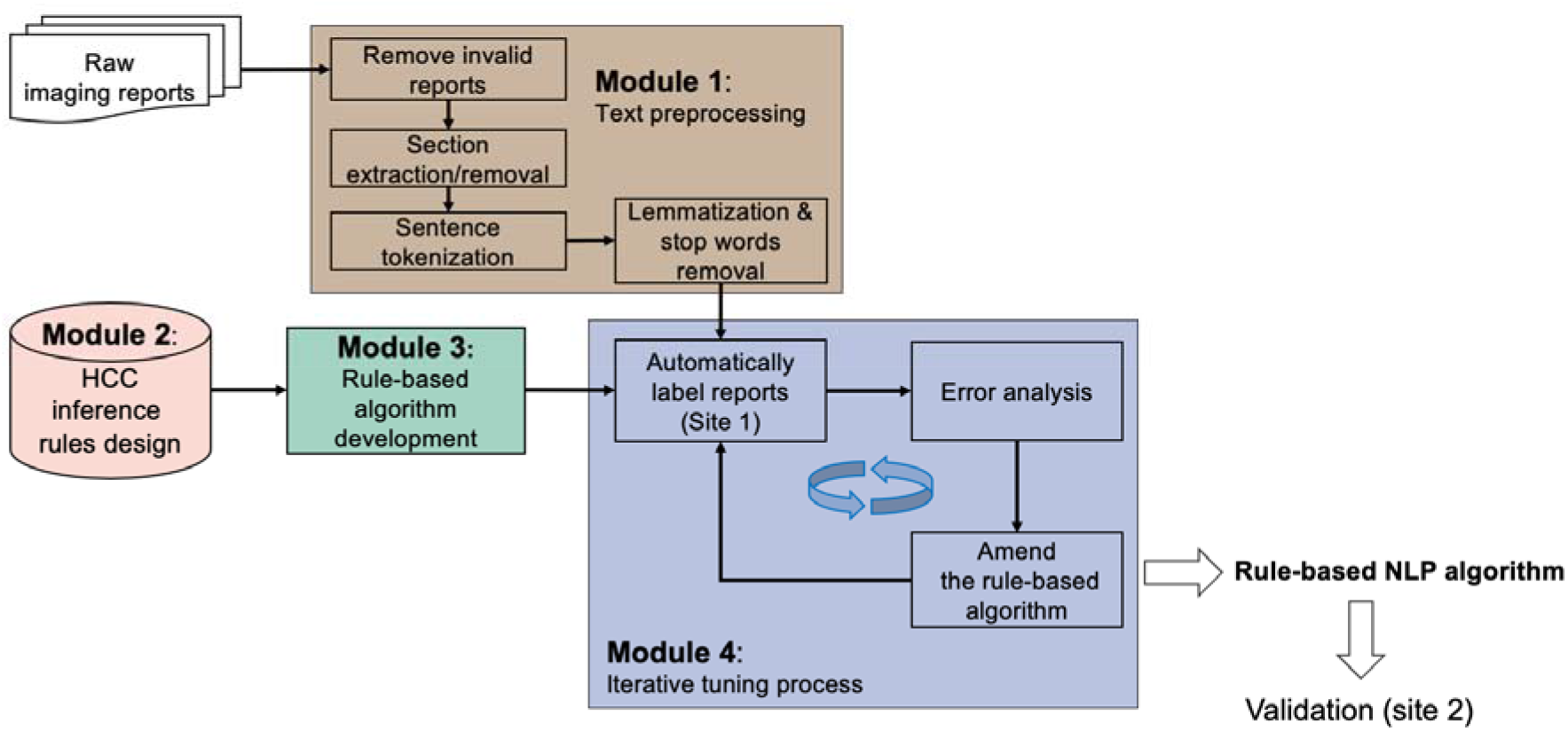
Framework of the proposed NLP method for identifying HCC from imaging reports

#### Module 1: Text preprocessing using NLP tools

All retrieved raw reports were preprocessed in Python 3.6 using NLP typical techniques, including Natural Language Toolkit (NLTK) and spaCy. Firstly, we excluded those invalid reports, e.g., with a “due to image omission or degradation” note, or blank reports. Then for the eligible reports, scripts were made to clean and pre-process the text (section extraction or removal). In this part, we applied the keywords matching algorithm to extract the impression part of an imaging report or remove the irrelevant description. Specifically, different strategies were used to tackle reports based on their characteristics (Figure S1):

- If an “impression” section was available in a report, we used this section for HCC identification. This was performed using regular expression for the words “impression”, “opinion”, “conclusion”, “summary”, “comment”, or their corresponding abbreviations or synonyms.
- For those reports without an “impression” section but with a “findings” section available, we extracted the “findings” section. Additionally, as the “findings” section typically includes descriptions of multiple body systems, we excluded the text irrelevant to liver system by removing sentences mentioning other body systems. To perform this process we built a lexicon of body systems that are not relevant to HCC information extraction, such as gallbladder, spleen, kidney, or their synonyms, and conditions related to these organs (eg. gallstone, splenomegaly, hydronephrosis) (Table S1).
- For those reports without an explicit “impression” or “findings” section title, we considered a removal of irrelevant content from the report. Specifically, as we are interested in the findings of the current imaging investigation, we excluded the sections such as “reason for study”, “reason for examination”, “clinical history”, or “clinical information”, by using keyword matching filters, and used the remaining text for a report.
- For those reports without any section titles, we split the report and excluded the text irrelevant to findings of liver system based on the built lexicon of other body systems as mentioned above.

For the extracted text, we then employed text processing techniques to reduce the complexity and noise of the data, and prepare the substantive content for further analysis. In particular, we first split reports into paragraphs using the new line character (as it not uncommon that imaging reports list relevant findings/impressions line by line) and then tokenise text into individual sentences. We also used lemmatization technique to standardise words and removed stop words (e.g., “an”, “of”, and “the”) that are used for connection and grammar but do not provide informative meaning. However, we did not consider words such as “nor”, “not”, “none”, “can’t”, “aren’t”, “amount” as stop words in this study, as they have negation meaning or useful information.

#### Module 2: Design of HCC inference rules for imaging reports

To build inference rules for HCC from imaging reports, key terms for the inclusion and exclusion criteria of HCC were highlighted by a senior hepatologist (E.B.) to form a list of “seed terms”. We then expanded the list based on multiple clinical guidelines for diagnosis of HCC based on imaging examination (7-9, 17). In the development phase, we initially ranked the priorities of the key terms to form diagnostic rules, with a reference to the LI-RADS (8). Typically, medical terms presenting in the “impression” section are different from those in the “findings” section in an imaging study report, as the former tend to be conclusive/diagnostic terms (e.g., “no evidence of hepatoma”) whilst the latter tend to be descriptive terms of organ/tumour features (e.g., “There are two subcentimetre foci of hypervascularity within segment 8 of the liver. There is no washout demonstrated”). Therefore, to identify HCC information from different sections we further classified key terms into two levels, one concerning concepts of tumour features and the other concerning the variations of tumour diagnostic concepts. The medical terms for HCC identification are provided in Table 1.

**Table 1.**
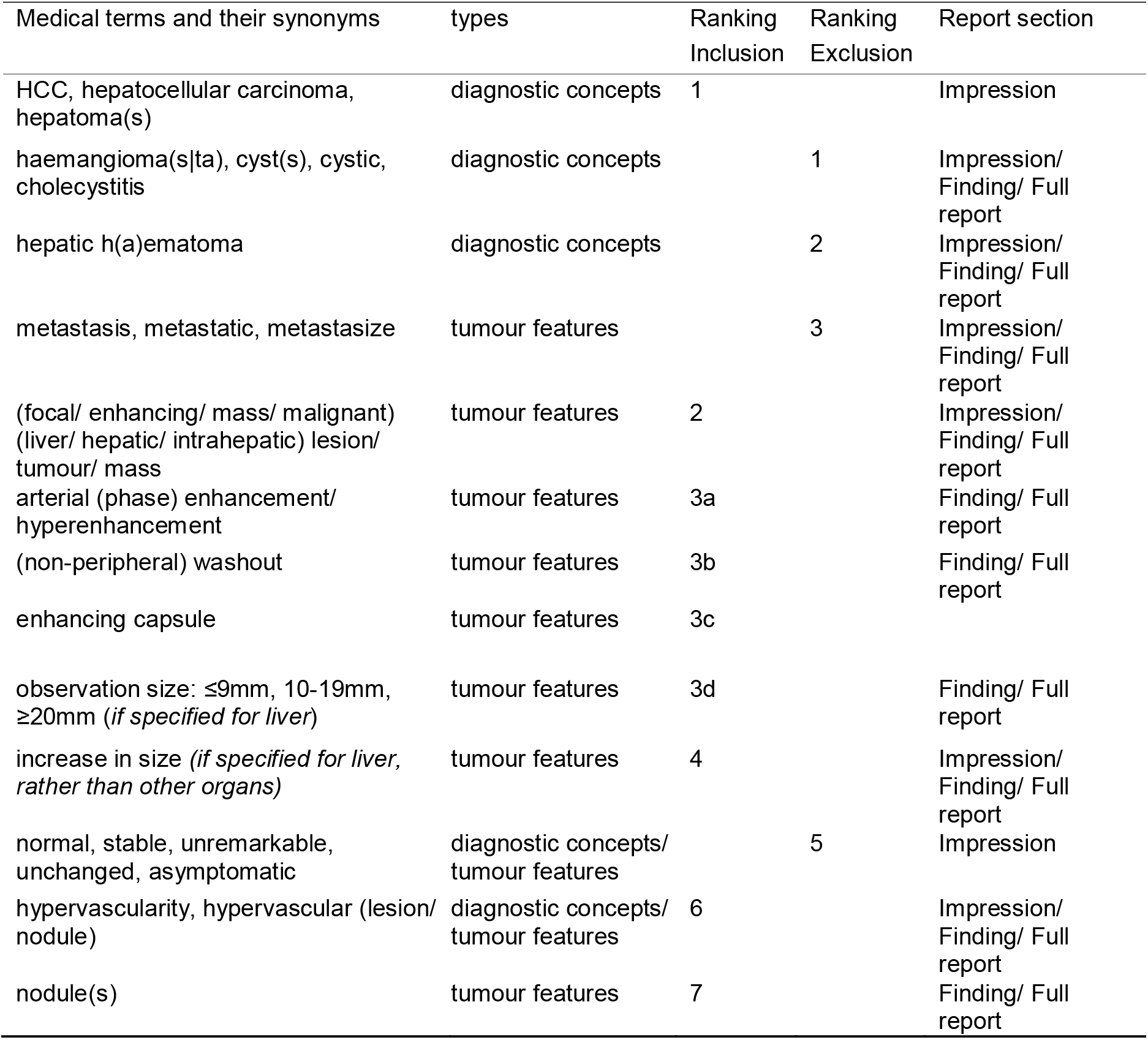
Medical terms and priority rankings in inference rules for hepatocellular carcinoma (HCC)

| Medical terms and their synonyms | types | Ranking Inclusion | Ranking Exclusion | Report section |
| --- | --- | --- | --- | --- |
| HCC, hepatocellular carcinoma, hepatoma(s) | diagnostic concepts | 1 |  | Impression |
| haemangioma(s/ta), cyst(s), cystic, cholecystitis | diagnostic concepts |  | 1 | Impression/<br>Finding/ Full report |
| hepatic h(a)ematoma | diagnostic concepts |  | 2 | Impression/<br>Finding/ Full report |
| metastasis, metastatic, metastasize | tumour features |  | 3 | Impression/<br>Finding/ Full report |
| (focal/ enhancing/ mass/ malignant) (liver/ hepatic/ intrahepatic) lesion/ tumour/ mass | tumour features | 2 |  | Impression/<br>Finding/ Full report |
| arterial (phase) enhancement/ hyperenhancement | tumour features | 3a |  | Finding/ Full report |
| (non-peripheral) washout | tumour features | 3b |  | Finding/ Full report |
| enhancing capsule | tumour features | 3c |  |  |
| observation size: ≤9mm, 10-19mm, ≥20mm (*if specified for liver*) | tumour features | 3d |  | Finding/ Full report |
| increase in size (*if specified for liver, rather than other organs*) | tumour features | 4 |  | Impression/<br>Finding/ Full report |
| normal, stable, unremarkable, unchanged, asymptomatic | diagnostic concepts/<br>tumour features |  | 5 | Impression |
| hypervascularity, hypervascular (lesion/ nodule) | diagnostic concepts/<br>tumour features | 6 |  | Impression/<br>Finding/ Full report |
| nodule(s) | tumour features | 7 |  | Finding/ Full report |

#### Module 3: Development of rule-based NLP algorithm for HCC identification

We used the initially-developed inference rules to design a rule-based NLP algorithm prototype. The logical process of the algorithm is to tag each sentence within a report and then aggregating all the tags to generate a final label for the report based on the inference rules set out (Figure S2). The algorithm module contains five submodules as follows:

- Regular expressions of medical terms. We iteratively built and tuned the regular expressions of target medical terms based on the development set (Table S2).
- Relevant medical terms recognition. We matched and extracted terms from imaging reports by the built regular expressions. To facilitate the following step of negation detection we replaced a matched term with a corresponding standard terminology that can be accurately recognised by a named entity recognition (NER) model.
- Negation detection. For detecting if a target term is negated in imaging reports, we built a negation detection module based on negSpacy (18) and Stanza. negSpacy is a pipeline developed based on NegEx (19) for identifying negations in text. Stanza is a collection of state-of-the-art NLP tools, for syntactic analysis and NER in both general and biomedical domains (20, 21). Specifically, our negation detection module firstly wraps Stanza, using the pre-trained syntactic analysis model based on Medical Information Mart for Intensive Care III (MIMIC-III) database and the pre-trained NER model based on MIMIC-III imaging reports. Then we added negSpacy components into our negation detection module. For negation patterns in negSpacy, we used the clinical sensitive version designed for clinical domain (18), and also added new customised negation patterns for imaging reports based on our development set.
- Sentence tagging based on priority (sentence-level). With identifying medical terms and detecting negation, we sequentially tagged a sentence based on the priority ranking to obtain a list of candidate tags for each sentence. We then assigned the tag with the highest priority among the candidate tags to each sentence. As a result, a set of sentence tags will be generated for each report.
- Inferring report label (document-level). To infer a final label (HCC or non-HCC) for each report, we sorted the sentence tags by the priority ranking, and generated report label using the sentence tag with highest priority.

#### Module 4: Iterative tuning process for the rule-based NLP algorithm

We used the development set to tune the designed rule-based NLP algorithm (Figure 1, Module 4). Specifically, we first utilised the algorithm to label reports, then checked the reason for errors so as to adjust the key terms lists and their priority ranking accordingly. The tuning process was iteratively performed until the accuracy could not be further improved.

### Evaluation metrics

To evaluate the algorithm performance, a confusion matrix was generated. The true positive (TP) value indicates the number of HCC-labelled reports that are correctly recognised by the algorithm as HCC, whilst the true negative (TN) value indicates the number of reports labelled with non-HCC and correctly recognised by the algorithm as non-HCC. The false positive (FP) value represents the number of reports labelled with non-HCC but wrongly recognised by the algorithm as HCC, whilst the false negative (FN) value represents the number of reports labelled with HCC but wrongly recognised by the algorithm as non-HCC. We used the following metrics to evaluate algorithm performance: sensitivity (aka, recall or true positive rate), TP/ (TP + FN); specificity (aka, true negative rate), TN/ (TN + FP); precision (aka, positive predictive value (PPV)), TP/ (TP + FP); accuracy, (TP + TN)/ (TP + FP + TN + FN), and; F1 score, TP/ (TP+1/2(FP+FN)). As the original labels of each report were binary, “probably positive” or “definitely positive” from our algorithm were collapsed as “positive”, and “probably negative” or “definitely negative” were collapsed as “negative”.

### ML and DL models for comparison

We compared the performance of our rule-based NLP algorithm against the following three categories of off-the-shelf supervised learning models: (a) conventional simple yet competitive ML based text classifiers, including Naive Bayes (NB), Logistic Regression (LR), and Support Vector Machine (SVM), with bag-of-words (BOW) or term frequency-inverse document frequency (TF-IDF) as the text representation methods (22). (b) DL based text classification architectures, including text convolutional neural network (CNN) (23) and hierarchical attention networks (HAN) (24), with word2vec as the representation method of text (25, 26). To obtain word embedding related to target domain of imaging reports, we trained a word2vec model on all reports of CT, MRI, or ultrasound scans from MIMIC-III database. (c) Fine-tuning pre-trained DL models with contextual representation of text, where we used Bidirectional Encoder Representations from Transformers (BERT) (27) and its variant versions in clinical domain including ClinicalBERT (28) and BlueBERT (29). For all the supervised learning models, we used data from site 1 (the development set) to train classifiers and evaluate the performance on data from site 2 (the test set) with the metrics above. As data are highly imbalanced, we applied cost-sensitive learning scheme during training to take into account the skewed distribution (30).

## RESULTS

### Descriptive characteristics of imaging reports

We included 1140 imaging reports with labels available (655 from site 1 and 485 from site 2) for this study. The earliest reports are sourced from the year 2003, while the latest reports are from the year 2019. Due to the long timespan (∼15 years) and reports collected from different sites, these imaging reports have variable reporting and linguistic styles. For both sites, the imaging modality of most reports was ultrasound, with CT and MRI examinations performed less frequently, in accordance with clinical guidelines (Table 2). For site 1 vs. site 2, the prevalence of HCC presence in ultrasound reports was 0.84% (5/598) vs. 1.27% (4/314), whilst 18.18% (6/33) vs. 5.43% (5/92) for CT reports and 12.50% (3/24) vs. 3.80% (3/79) for MRI reports.

**Table 2.** Distribution of imaging modalities and HCC prevalence (labels annotated by medical students) in each site

| Imaging modalities | Site 1 (n=655) |  | Site 2 (n=485) |  |
| --- | --- | --- | --- | --- |
|  | Total | Labelled with HCC | Total | Labelled with HCC |
| Ultrasound | 598 | 5 (0.84%) | 314 | 4 (1.27%) |
| CT | 33 | 6 (18.18%) | 92 | 5 (5.43%) |
| MRI | 24 | 3 (12.50%) | 79 | 3 (3.80%) |

CT, computed tomography; HCC, hepatocellular carcinoma; ICHT, Imperial College Healthcare NHS Trust; MRI, magnetic resonance imaging.

Text characteristics of reports across the two sites are summarised in Table 3. The proportion of reports that had “impression” sections was varied between sites. Only 18% (117/655) of reports from site 1 included an “impression” section, while the remaining 82% of reports did not, making HCC identification more challenging. In contrast, 75% (366/485) of the reports from site 2 had “impression” sections. The number of sentences or words per report was significantly different between the two sites, both before and after text pre-processing (all p values <0.001), which reflects significant heterogeneity between sites. The information of running time for data preprocessing are provided in Table S3.

**Table 3.**
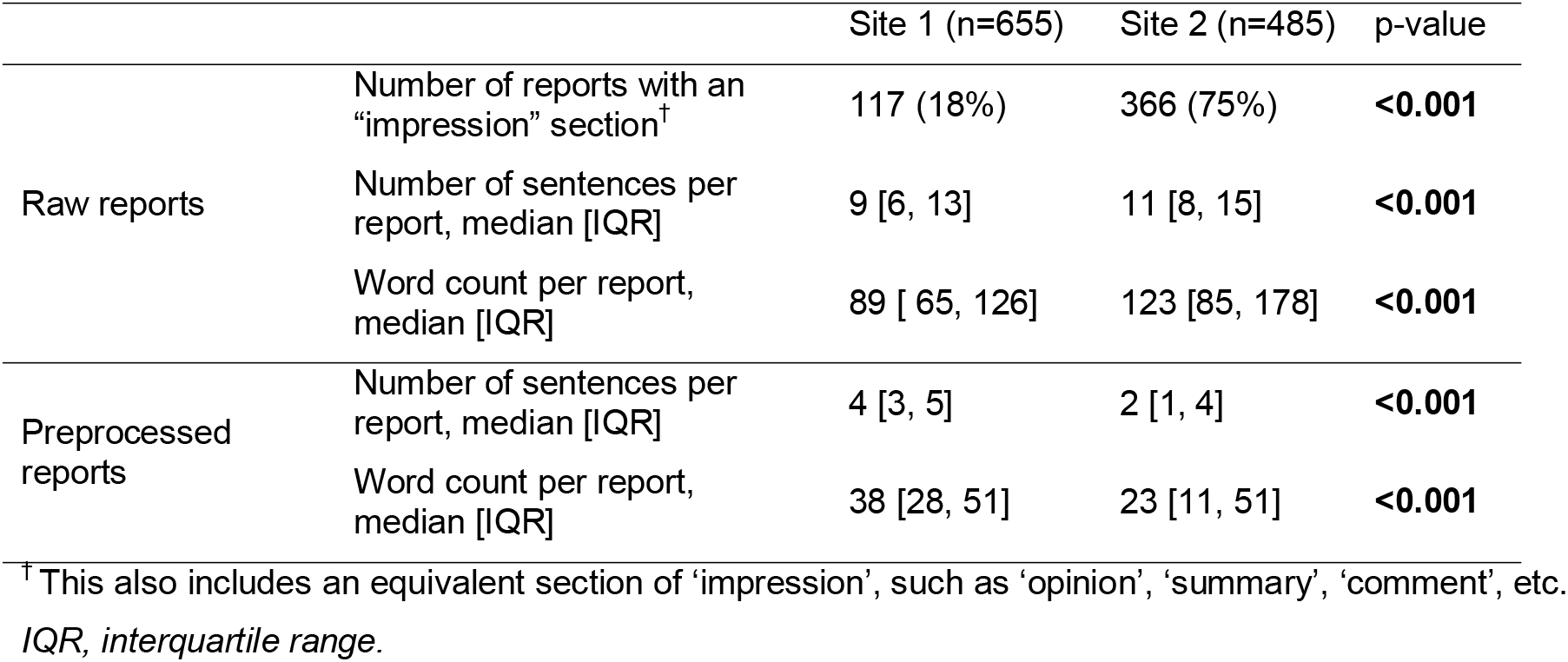
Text characteristics of imaging reports across two sites

### Performance on the development set

After iteratively tuning and amending the algorithm using the development set, the priority ranking of terms in diagnostic rules were finalised (Table 1). Compared to the original labels in the development set, our algorithm obtained an overall consistency of 99.38%, with 71.43% consistency on HCC cases and 100% on non-HCC cases, with a short duration of running time (Table S3). The confusion matrix is shown in Figure 2A. However, among the five HCC-labelled reports being identified as non-HCC by the algorithm, four were actually non-HCC reports after further manual checking, and the remaining one was incorrectly identified by the algorithm (Table S4). To further verify the correctness of our algorithm to label non-HCC cases, we randomly selected 10% (64/641) of original non-HCC reports by stratified sampling (59 for ultrasound, 3 for CT, 2 for MRI), and manually checked the ground truth labels of these reports. We found that all the 64 reports were accurately identified by the algorithm. We then generated a new confusion matrix after data correction (Figure 2B), and the overall accuracy is 99.85%, with 90% (9/10) of sensitivity and 100% (645/645) of specificity.

**Figure 2.**
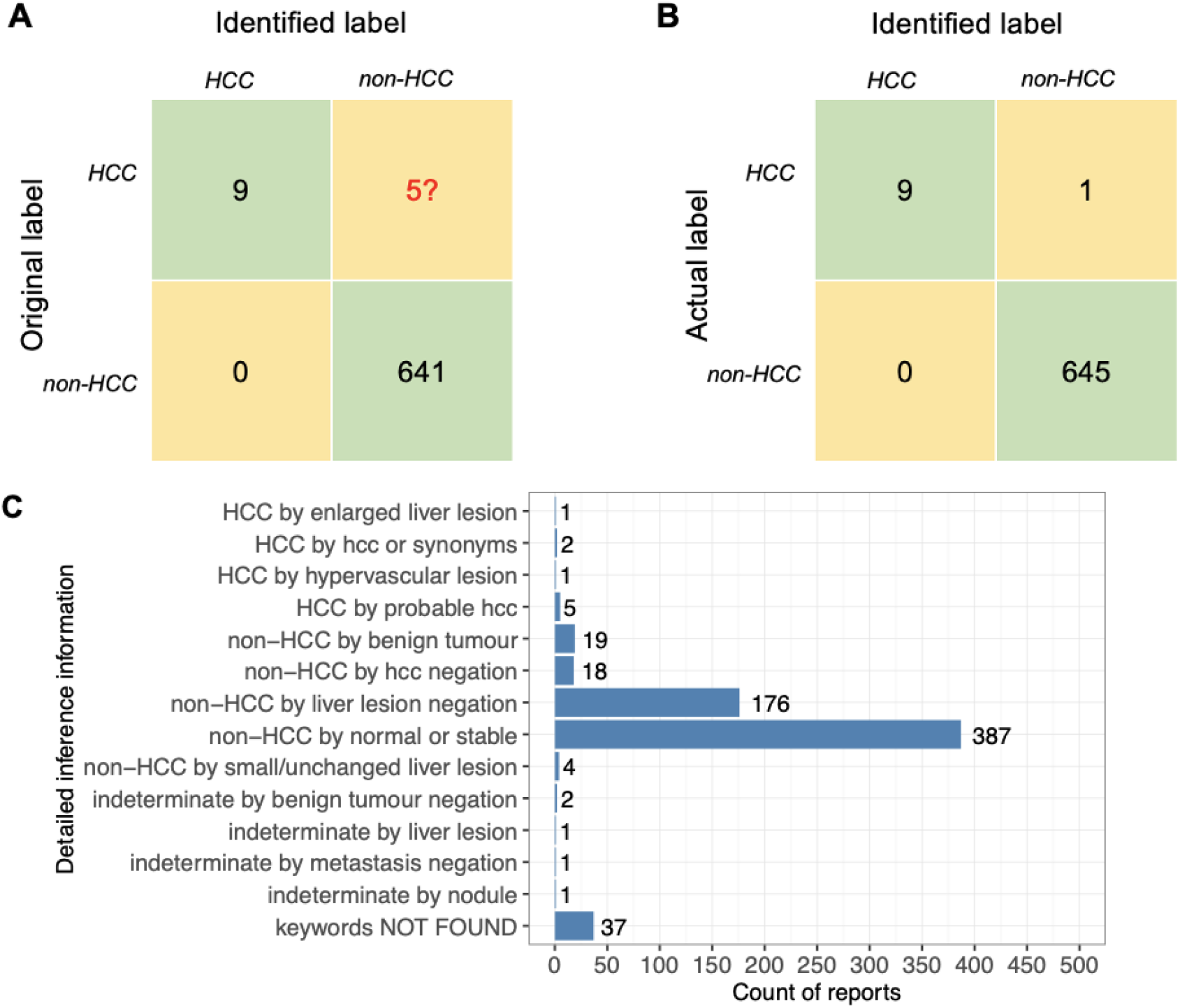
Results in development set (site 1): (A) confusion matrix compared to raw labels; (B) confusion matrix compared to actual (ground-truth) labels; (C) Distribution of each criteria used by the algorithm in development set. *Noting that the categories of ‘keywords not found’ or ‘indeterminate’ were classified into ‘non-HCC’ by the algorithm*.

We summarised the frequency of terms used for the identification process in the development set (Figure 2C). HCC presence identification was derived mostly based on “(probable) HCC” or its synonyms (“hepatoma/ hepatocellular carcinoma”), followed by “enlarged liver lesion” or “hypervascular lesion”. In contrast, non-HCC (HCC absence) identification was inferred mostly based on normal examination or negation of liver lesion, followed by benign tumour or negation of HCC.

### Performance on the validation set

Compared to the originally collected labels in the test set, the algorithm achieved an overall consistency of 99.18% (481/485), with 91.67% consistency (11/12) on HCC cases, and 99.37% (470/473) on non-HCC cases; the confusion matrix is shown in Figure 3A. However, for the one HCC-labelled report being identified as non-HCC, the ground-truth label of the report is in fact “non-HCC” (Table S5). For the three non-HCC-labelled reports being identified as HCC, one report was in fact highly suspicious of HCC, two reports were equivocal or indeterminate (Table S5). To further verify the correctness of our proposed algorithm on non-HCC cases in the test set, 47 reports (10%) of the original 468 non-HCC reports were randomly selected, for which ground truth labels were manually checked. We found that all these 47 reports were accurately identified by our proposed NLP algorithm.

**Figure 3.**
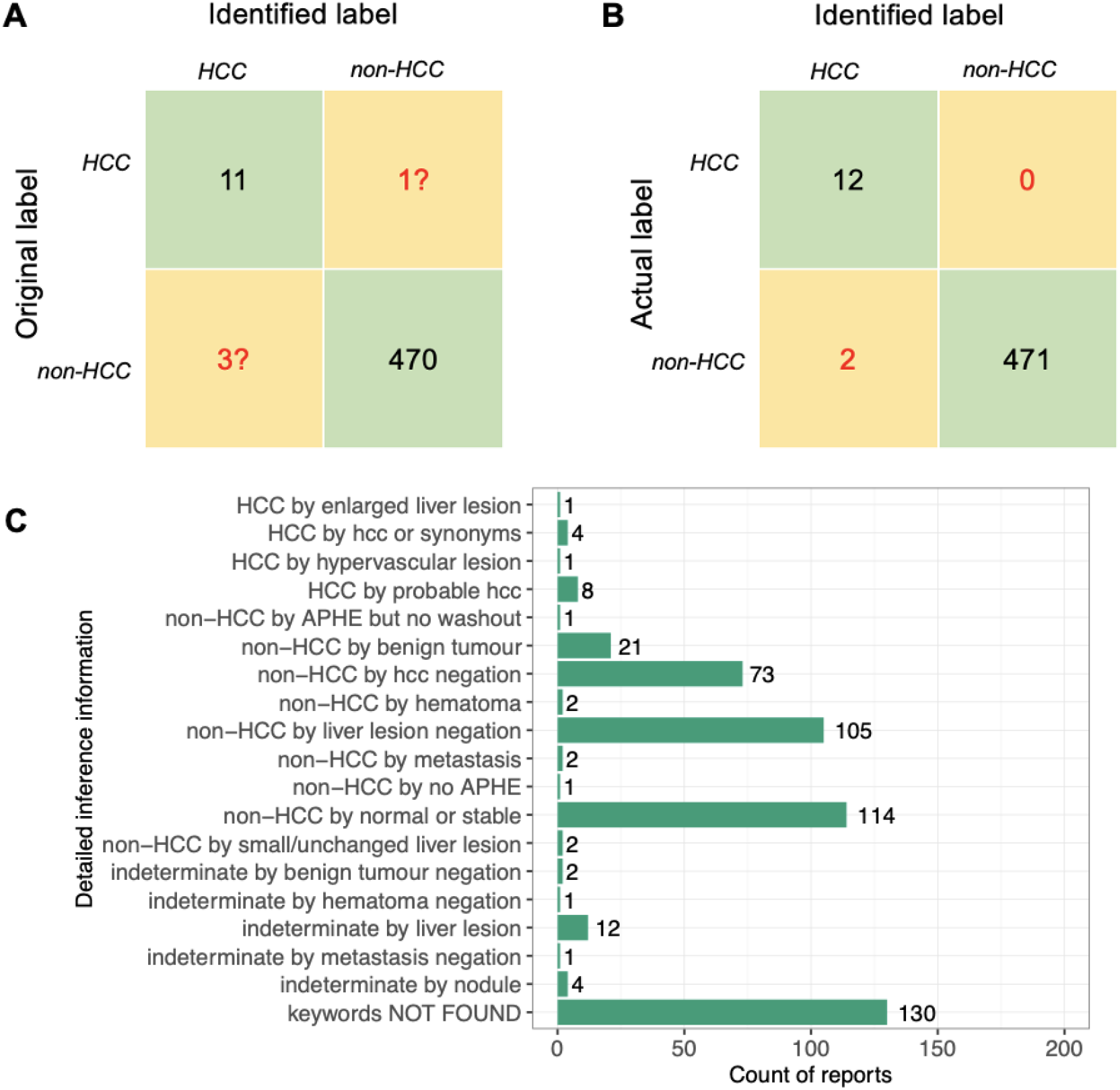
Results in test set (site 2): (A) Normalised confusion matrix compared to raw labels, (B) Normalised confusion matrix compared to ground-truth labels, (C) Distribution of each criteria used by the algorithm in test set. *Noting that the categories of ‘keywords not found’ or ‘indeterminate’ were classified into ‘non-HCC’ by the algorithm*.

After data correction, the overall accuracy of the rule-based NLP method on the test set is 99.59% (483/485), with 100% sensitivity and 99.58% (471/473) specificity (Figure 3B).

Similar to that of the development set, we also summarised the frequency of terms used for identifying the process in the test set (Figure 3C). HCC presence identification was derived mostly based on “(probable) HCC” or its synonyms (“hepatoma/ hepatocellular carcinoma”), followed by “liver lesion” and “hypervascular lesion”. In contrast, non-HCC (HCC absence) identification was derived mostly based on normal examination or negation of liver lesion, then followed by negation of HCC.

### Comparison of performance between our rule-based method and ML/DL models

We used the development set (after data correction) to train ML and DL based models. The performance comparisons between the proposed rule-based NLP method and supervised learning models in the test set are summarised in Table 4. The rule-based method was superior to conventional ML models regardless of the feature representation method (BOW or TF-IDF) as well as outperformed DL models (CNN, HAN, BERT, ClinicalBERT, or BlueBERT). Particularly, the rule-based method achieved significantly higher sensitivity compared to other models, although the rule-based method demonstrated only a marginal improvement in overall accuracy due to a small number of HCC cases. HAN with word2vec obtained a higher sensitivity (75%) than other supervised learning models; however, it is still significantly inferior to the rule-based method (100%). Although the pre-trained transformers models can achieve comparable specificity (97.67% to 99.79%), they had low sensitivity (33.33% to 58.33%) due to small sample size of data for the class of HCC. The rule-based method was capable of handling skewed data distributions by achieving a much higher F1 score and prevision (F1 score = 0.9231; Precision = 99.59%) compared to supervised learning models regardless of model architecture and feature representation method (F1 score = 0.11 to 0.47; Precision = 6.3% to 80.0%).

**Table 4.**
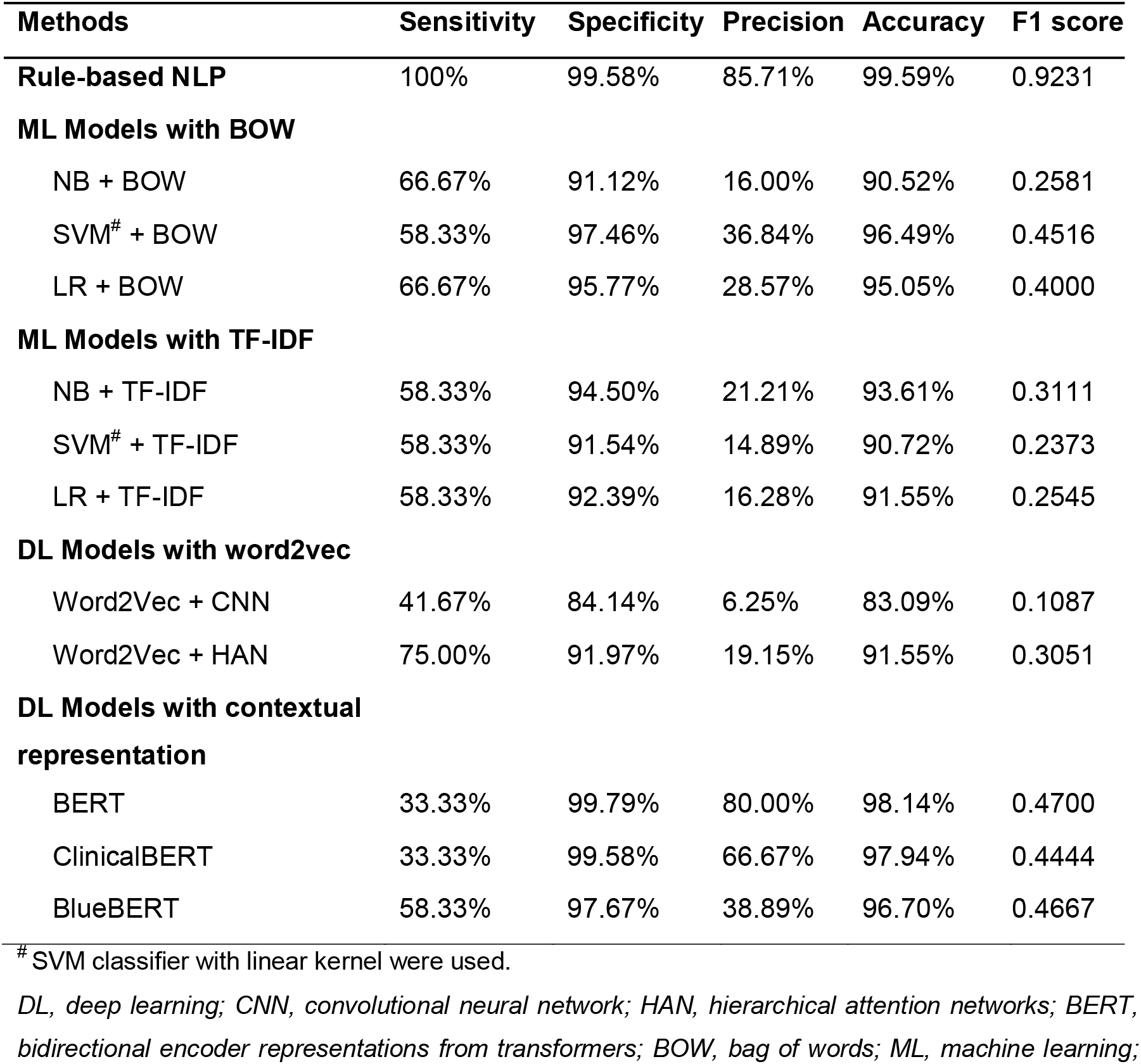
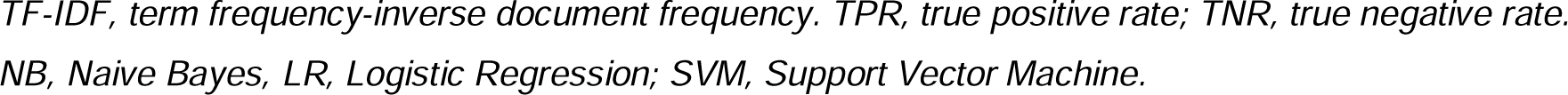
The performance comparison between the proposed rule-based method and ML- or DL-based models in the test set (Site 2) after label correction

| Methods | Sensitivity | Specificity | Precision | Accuracy | F1 score |
| --- | --- | --- | --- | --- | --- |
| **Rule-based NLP** | 100% | 99.58% | 85.71% | 99.59% | 0.9231 |
| **ML Models with BOW** |  |  |  |  |  |
| NB + BOW | 66.67% | 91.12% | 16.00% | 90.52% | 0.2581 |
| SVM<sup>#</sup> + BOW | 58.33% | 97.46% | 36.84% | 96.49% | 0.4516 |
| LR + BOW | 66.67% | 95.77% | 28.57% | 95.05% | 0.4000 |
| **ML Models with TF-IDF** |  |  |  |  |  |
| NB + TF-IDF | 58.33% | 94.50% | 21.21% | 93.61% | 0.3111 |
| SVM<sup>#</sup> + TF-IDF | 58.33% | 91.54% | 14.89% | 90.72% | 0.2373 |
| LR + TF-IDF | 58.33% | 92.39% | 16.28% | 91.55% | 0.2545 |
| **DL Models with word2vec** |  |  |  |  |  |
| Word2Vec + CNN | 41.67% | 84.14% | 6.25% | 83.09% | 0.1087 |
| Word2Vec + HAN | 75.00% | 91.97% | 19.15% | 91.55% | 0.3051 |
| **DL Models with contextual representation** |  |  |  |  |  |
| BERT | 33.33% | 99.79% | 80.00% | 98.14% | 0.4700 |
| ClinicalBERT | 33.33% | 99.58% | 66.67% | 97.94% | 0.4444 |
| BlueBERT | 58.33% | 97.67% | 38.89% | 96.70% | 0.4667 |

<sup>#</sup> SVM classifier with linear kernel were used.
*DL, deep learning; CNN, convolutional neural network; HAN, hierarchical attention networks; BERT, bidirectional encoder representations from transformers; BOW, bag of words; ML, machine learning;*
TF-IDF, term frequency-inverse document frequency. TPR, true positive rate; TNR, true negative rate. NB, Naive Bayes, LR, Logistic Regression; SVM, Support Vector Machine.

## DISCUSSION

In this study, we designed a rule-based NLP algorithm to identify the presence of HCC from free-text imaging reports. With a reference to ground-truth labels, the proposed rule-based NLP method achieve superior performance, especially higher sensitivity in highly-imbalanced datasets with small numbers of positive cases, compared to existing off-the-shelf models. The inner-working and sensitivity of the proposed method are not related to imaging modality, because we considered the major features used for HCC identification in different imaging modalities. For those labelled reports, the proposed method can be used to correct data, as we found some reports were incorrectly labelled during data collection, especially for HCC presence. For those unlabelled reports, the proposed method is capable of rapidly identifying the presence of HCC from the imaging reports, so as to provide labels for further cohort studies to advance HCC-related research.

Rule-based methods, ML or DL based models, or hybrid methods have been used to develop NLP pipelines in imaging reports (31-36), but few studies have focused on HCC identification (13, 14). A previous study has applied a hybrid NLP algorithm (Automated Retrieval Console, ARC), that was originally designed for other cancers, in HCC identification from free-text medical notes (13), wherein HCC were firstly identified by ICD-9 codes and then further verified by the hybrid NLP algorithm. With a pre-selection of candidate reports by ICD codes, this method can avoid the imbalanced data for text classification, whilst the issue of HCC misclassification by ICD codes is not considered. As LI-RADS has been well applied in US, another study (14) trained ML based models for HCC identification, which used imaging reports created from a LI-RADS template as labelled data. However, it is difficult to transfer such methods to those regions where LI-RADS standard are not widely used. Our study differs from previous studies and is novel in that we designed a rule-based NLP which utilised both tumour feature and diagnostic concept representations of HCC, along with the priority ranking of these terms, for which we accounted for linguistic styles in different sections of reports. Therefore, our algorithm has high specificity and perfect sensitivity in the datasets we have interrogated, and is capable of accurately identifying the presence of HCC, even in a small and highly-imbalanced dataset.

Two types of methods are typically used to handle the issue of highly imbalanced classes in text classification: a) resampling method and b) cost-sensitive learning scheme (30). In our study, the sample size of the minority class (HCC) was too small to apply resampling methods (specifically oversampling techniques such as SMOTE), which may produce un-representative synthetic examples for the minority class (30, 37). Therefore, we used a cost-sensitive learning scheme, putting greater weights on the minority class (in this case, HCC) to tackle the issue of imbalance. However, even in our study the ML or DL models trained with this imbalanced learning scheme, were unable to achieve comparable performance to the proposed rule-based method, due to limited data on positive cases for supervised learning.

Compared to ML or DL models, the advantages of a rule-based method are that a large amount of labelled data is not required for development and validation, and that the method is useful as a cold-start to automatically label reports. It is therefore sensible to choose a rule-based method at the initial phase, instead of ML or DL models as large volume of labelled data is needed for training such models to achieve high performance, particularly when a large imbalance between positive and negative cases exists.

Our rule-based NLP method achieved a high accuracy on both HCC and non-HCC cases and it is capable of rapidly labelling data on a large-scale. Our findings provide an opportunity for DL to be applied in HCC identification in future by weak supervision schemes (38, 39). Weak supervision uses noisy labelled data that might include incorrect labels to train supervised learning models. DL has been demonstrated to be valuable and robust in medical text mining tasks, even in noisy data. However, the lack of labelled data is a major bottleneck to applying such technique for disease information extraction (40). Recently DL-based NLP models trained with rule-based method labelled data (weak supervision learning) has been successfully applied into medical free-text mining tasks such as hip fracture classification (38) and Alzheimer’s disease risk factor characterisation(41), achieving competitive performance compared to models trained on human-annotated data. Promisingly, our proposed rule-based NLP method provides a foundation for application of these techniques to identify HCC from free-text imaging reports. Data collection within the NIHR HIC viral hepatitis theme is still ongoing (15), and therefore it is expected that a large amount of unlabelled imaging reports will be collected from more collaborators across England in the near future. Reports can be automatically labelled by our rule-based NLP pipeline and accordingly can be fed into a DL model for training.

It is well recognised that a rule-based method typically has limited flexibility and robustness. Misspellings and numerous non-standardised ways of describing the same concept are common in imaging reports. Rule-based NLP systems may not work well if vocabularies and linguistic styles are significantly different. In the future, with more data accumulated, a more robust and flexible NLP pipeline using DL will be developed on the basis of the proposed rule-based method as discussed above. In addition, staging HCC is important to determine outcomes and planning of optimal therapy (17). Future work will also extend the proposed methodology for HCC staging.

## Conclusion

A rule-based NLP method to classify free-text imaging reports is developed to accurately identify HCC information for highly-imbalanced small-size dataset. When labelled data are not sufficiently available, a rule-based NLP algorithm with an integration of the domain knowledge can achieve a desirable accuracy, compared to supervised learning algorithms. Such a rule-based NLP algorithm not only can be utilised to rapidly screen patient free-text imaging reports on a large scale to facilitate epidemiological and clinical studies but also can serve as a tool for data correction.

## Data Availability

Data from NIHR HIC viral hepatitis theme may be made available to researchers on request following positive review by the steering committee. Further details are available at https://hic.nihr.ac.uk. Queries regarding data access should be directed to. MIMIC is provided through the work of researcher at the MIT Laboratory for Computational Physiology and the collaborators. Data are available through formally requesting access with the steps here: https://mimic.mit.edu/docs/gettingstarted/.

https://mimic.mit.edu/docs/gettingstarted/

## Ethics approval and consent to participate

The research database for the NIHR HIC viral hepatitis theme was approved by South Central - Oxford C Research Ethics Committee (REF Number: 15/SC/0523). All methods in this study were carried out in according to relevant guidelines and regulations. The requirement for written informed consent was waived by South Central - Oxford C Research Ethics Committee, because data have been anonymised before its use and the study is retrospective.

## Acknowledgements

The authors would like to thank all the research nurses and research admin staff at the contributing site for their help in data collection and submission.

## Author contributions

TW, EB, GSC, and JD conceived the study. BG, LM, DP, CRJ, TW, DAS, HS, CC, OF, SH, KAV, GR, SL, TN, and KW contributed to the acquisition, processing, interpretation, or management of data from study sites. TW designed the analytical strategy and conducted the analysis, supervised by EB, GSC, and JD. EB, GSC, and PCM helped interpret the findings. TW drafted the original manuscript with input from EB. All authors reviewed and revised the manuscript critically and approved the final version for publication.

## Code availability

The codes of the rule-based algorithm implementation will be made publicly available at github repository for this study: https://github.com/tinaty/NLPHCC.

## Abbreviations

AASLD: American Association for the Study of Liver Diseases
ACR: American College of Radiology
AFP: Alpha fetoprotein
APASL: Asian Pacific Association for the Study of the Liver
BERT: bidirectional encoder representations from transformers
BOW: bag-of-words
CNN: convolutional neural network
CT: multiphasic computed tomography
DL: deep learning
EASL: European Association for the Study of the Liver
EHR: electronic health record
ESMO: European Society for Medical Oncology
FP: false positive
FN: false negative
HAN: hierarchical attention networks
HBV: hepatitis B virus
HCC: hepatocellular carcinoma
HCV: hepatitis C virus
HIC: Health Informatics Collaborative
ICHT: Imperial College Healthcare NHS Trust
LI-RADS: liver imaging reporting and data system
LR: logistic regression
MRI: magnetic resonance imaging
NB: Naive Bayes
NHS: National Health Service
NIHR: National Institute for Health Research
NLP: natural language processing
NLTK: Natural Language Toolkit
OUH: Oxford University Hospitals
SVM: support vector machine
TF-IDF: term frequency inverse document frequency
TP: true positive
TN: true negative.

