## Supplementary Material for "Identifying Hepatocellular Carcinoma from imaging reports using natural language processing to facilitate data extraction from electronic patient records"

**Table S1**. Lexicon of body systems and conditions that are described in liver and abdomen imaging reports but are irrelevant to findings of hepatocellular carcinoma

| **Body systems and conditions** | **Conditions** |
| --- | --- |
| Gallbladder | Gallstone |
| Spleen | Splenomegaly |
| Kidney/renal | Hydronephrosis |
| Lung |  |
| Lumbar spine |  |
| Pancreas |  |
| Pelvis |  |
| Urinary bladder |  |
| Common bile duct or CBD |  |
| Biliary tree |  |

**Table S2. Regular expressions of medical terms of interest**

| **Medical terms** | **Regular expressions** |
| --- | --- |
| HCC  (along with modifiers) | ['hcc'] = re.compile(r"((?<=^)\|(?<= ))(hcc\|hepatocellular carcinoma\|hepatomas?)(?=\W\|$)", re.I \| re.M)  ['modifier_probable'] = re.compile(r"(probable\|suspicious\|likely\|possible\|may)") |
| Benign tumours | ["hemangiomas"] = re.compile(r"((?<=^)\|(?<= ))(ha?emangioma(ta\|s)?)(?=\W\|$)" , re.I \| re.M)  ["cyst"] = re.compile(r"((?<=^)\|(?<= ))(cyst(s\|ic)?)(?=\W\|$)" , re.I \| re.M)  ["cholecystitis"] = re.compile(r"((?<=^)\|(?<= ))(cholecystitis)(?=\W\|$)" , re.I \| re.M) |
| Haematoma | re.compile(r"((?<=^)\|(?<= ))(ha?ematoma)(?=\W\|$)", re.I \| re.M) |
| Metastasis | re.compile(r"((?<=^)\|(?<= ))(metastas(i\|e)s\|metastatic\|metastasize)(?=\W\|$)", re.I \| re.M) |
| Liver lesion/tumour  (along with modifiers) | re.compile(r"((?<=^)\|(?<= ))((focal\s\|enhancing\s\|mass\s)?(liver\s)?(lesions?\|tumours?\|mass(es)?))(?=\W\|$)" , re.I \| re.M)  ['modifier_enlarge'] = re.compile(r"[^(no)] enlarged?\|large\|increased?", re.I \| re.M)  ['modifier_unchanged'] = re.compile(r"not? changed?\|not? changes?\|unchanged?\|unchanges?\|stable\|no enlarges?\|no enlarged?", re.I \| re.M)  ['modifier_small'] = re.compile(r"small\|tiny\|little", re.I \| re.M) |
| APHE | re.compile(r"((?<=^)\|(?<= ))(((hyper)?enhancement.*\sarterial(\sphase)?)\|(arterial(\sphase)?.*\s(hyper)?enhancement)\|APHE\|(arterial(\sphase)? hypervascular(ity)?))(?=\W\|$)", re.I \| re.M) |
| Washout | re.compile(r"((?<=^)\|(?<= ))((non(-\| )?peripheral )?washout)(?=\W\|$)", re.I \| re.M) |
| Capsule  (along with modifiers) | ['capsule'] = re.compile(r"((?<=^)\|(?<= ))(((non-?)?enhancing )?capsule)(?=\W\|$)", re.I \| re.M)  ['modifier_enhancing'] = re.compile(r"((?<=^)\|(?<= ))enhancing(?= capsule)", re.I \| re.M)  ['modifier_nonenhancing'] = re.compile(r"((?<=^)\|(?<= ))non(-\| )?enhancing(?= capsule)", re.I \| re.M) |
| observation size | re.compile(r"((?<=^)\|(?<= ))(liver\|mass\|tumour\|lesion\|nodule)(\s\|\w)+(\d)+(\.\d+)?(\s)?(cm\|mm)(?=\W\|$)", re.I \| re.M) |
| Size increase | re.compile( r"((?<=^)\|(?<= ))(liver.+(increase in size))\|((increase in size).+liver)(?=\W\|$)", re.I \| re.M) |
| Normal examination | re.compile( r"((?<=^)\|(?<= ))(normal\|stable\|unremarkable\|unchanged\|asymptomatic)(?=\W\|$)", re.I \| re.M) |
| Hypervascular lesion | re.compile(r"((?<=^)\|(?<= ))(hypervascular(ity)?( lesion(s)?\| nodule(s)?)?)(?=\W\|$)", re.I \| re.M) |
| Nodules | re.compile(r"((?<=^)\|(?<= ))(nodules?)(?=\W\|$)", re.I \| re.M) |

**Table S3. Running time* of data preprocessing and report labelling by the proposed rule-based model for the development set and validation set**

| Dataset | Total running time for data preprocessing | Total running time for report labelling |
| --- | --- | --- |
| Development set (Site 1, 655 reports) | 0.18 mins | 1.77 mins |
| Validation set (Site 2, 485 reports) | 0.10 mins | 1.25 mins |

** The programs were running on a computing environment with CPU memory of 32 GB.*

**Table S4**. Inconsistent identifications (compared to original labels) and reasons for site 1

| Reasons resulting in inconsistent classification | Note |
| --- | --- |
| Reasons for five HCC-labelled reports being identified as non-HCC:  1 report: no obvious focal lesion  1 report: no mass lesions  1 report: liver is normal in size, no focal mass  1 report: possible haemangioma  1 report: wrong identification by the algorithm (liver lesion in segment VIII has enlarged slightly, no evidence of distant metastases) | data error  data error  data error  data error  false negative |

**Table S5**. Inconsistent identifications (compared to original labels) and reasons for site 2

| Reasons resulting in inconsistent classification | Note |
| --- | --- |
| Reasons for one HCC-labelled report being identified as non-HCC:  1 report: hepatic haematoma, which is non-HCC case | data error |
| Reasons for three non-HCC-labelled reports being identified as HCC:  1 report: highly suspicious of a diffuse hepatocellular carcinoma  1 report: equivocal (a questionable 2.6 cm high density lesion, may be an HCC or regenerative nodule)  1 report: equivocal (may represent regenerating nodules or haemangiomata but focal hepatomas cannot be excluded) | data error  equivocal  equivocal |

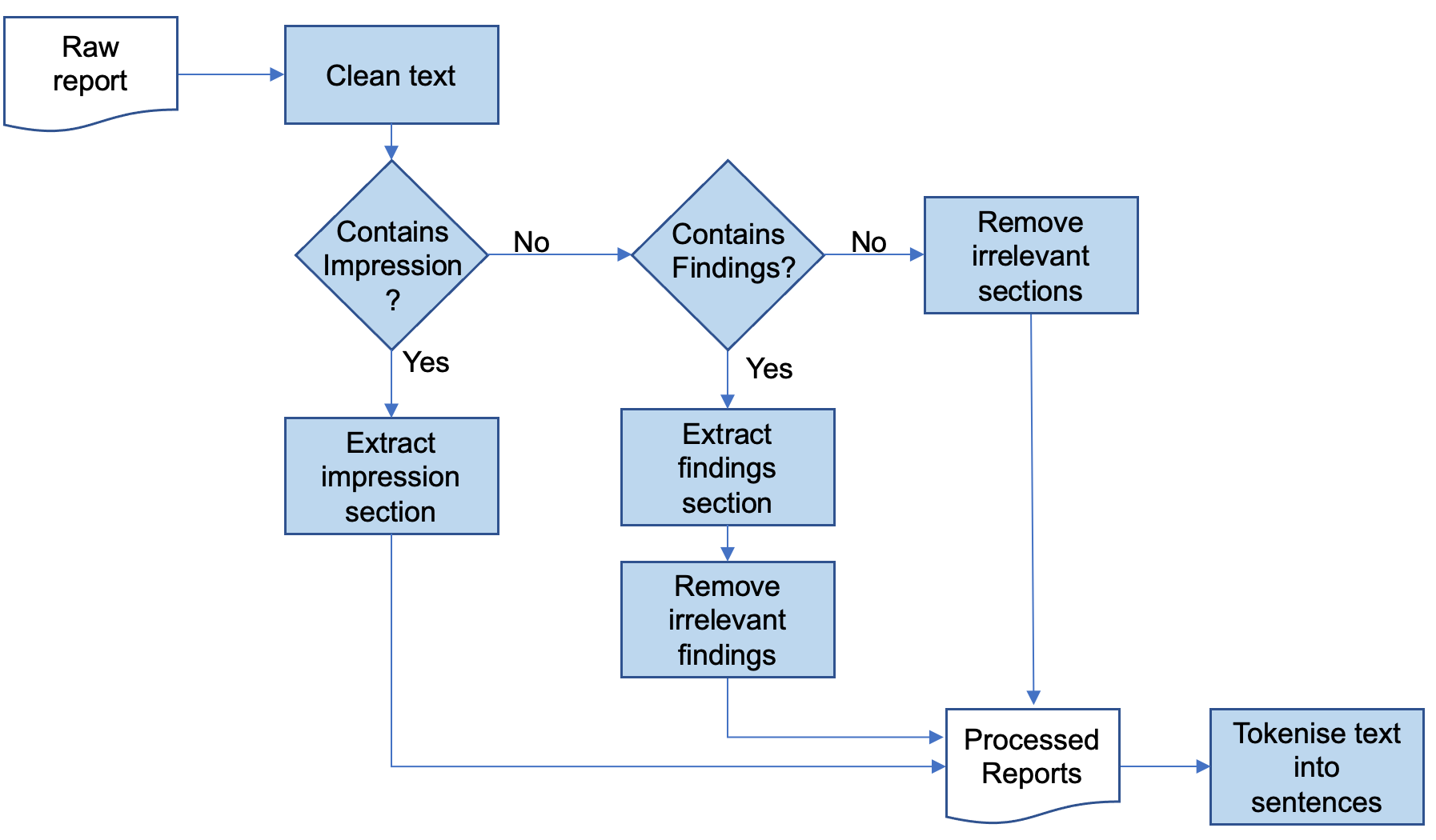

**Figure S1**. The text pre-processing pipeline for imaging reports

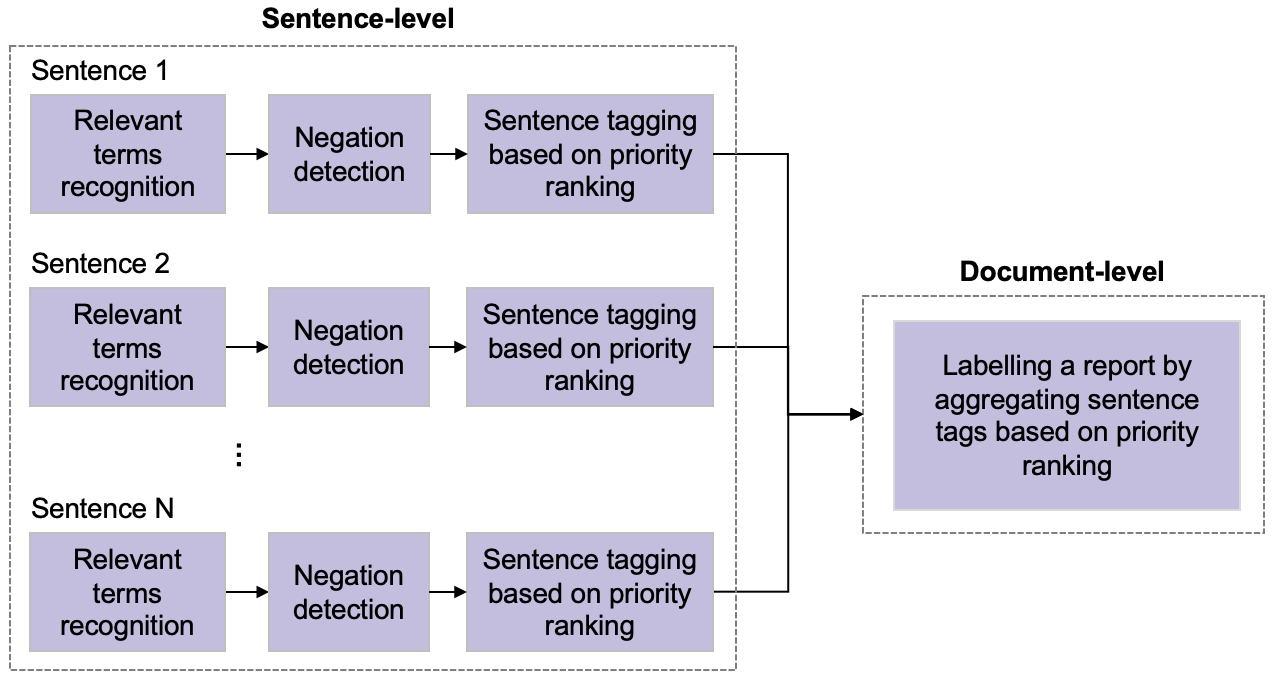

**Figure S2**. The logical process of labelling a report by the proposed rule-based NLP algorithm
